# Estimating Subnational Excess Mortality in Times of Pandemic. An application to French *départements* in 2020

**DOI:** 10.1101/2022.12.12.22283346

**Authors:** Florian Bonnet, Carlo Giovanni Camarda

## Abstract

The Covid-19 pandemic did not affect sub-national regions in a uniform way. Knowledge of the impact of the pandemic on mortality at the local level is therefore an important issue for better assessing its burden. Vital statistics are now available for an increasing number of countries for 2020 and 2021, and allow the calculation of sub-national excess mortality. However, this calculation faces two important methodological challenges: (1) it requires appropriate mortality projection models; (2) small populations implies important uncertainty in the estimates, commonly neglected. We address both issues by adopting a method to forecast mortality at sub-national level and by incorporating uncertainty in the computation of mortality measures. We illustrate our approach to French *départements* (NUTS 3, 95 geographical units) and produce estimates for 2020 and both sexes. Nonetheless, the proposed approach is so flexibility to allow estimation of excess mortality during Covid-19 in most demographic scenarios as well as for past pandemics.

## 1 Introduction

Since the emergence of the disease, estimating mortality due to Covid-19 has been the object of intense research, both to guide public policies aimed at controlling the circulation of the virus, and to know the global burden of this pandemic in various countries. National health surveillance agencies were first mobilized to provide weekly or even daily death tolls attributable to Covid-19, and thus establish a rapid indicator of the impact of the pandemic [1; 2]. However, differences in data definitions among countries, time-varying collection methods, delays in reporting and diverse coverage by place of death are known issues that hinder health surveillance systems to be employed for a reliable assessment of the pandemic [3; 4].

Over the months, the regular systems of official statistics have provided information that complements and/or corrects data provided by surveillance systems, including deaths by age from all causes. These data are the basis for the construction excess mortality measures as alternative method for assessing the burden of the pandemic. Defined as “the difference between the number of deaths (from any cause) that occur during the pandemic and the number of deaths that would have occurred in the absence of the pandemic” [5, p. 85], excess mortality can also be computed on other indicators like life expectancy and standardized death rates. Measures of excess mortality have been considered the gold standard to estimate the impact of Covid-19 [6; 7] and they have been extensively adopted in the past years. Whereas some of them used pre-pandemic years as baseline mortality in the absence of Covid-19 [8; 9], others have accounted for mortality changes over time [10; 11; 12; 13; 14].

However all of these studies estimate excess mortality at the national level. This perspective generally hides large regional differences that ought to be taken into account to better inform policy makers. Hence, in recent months, many papers have attempted to estimate excess mortality at a regional level. For some of these studies, excess mortality was the difference between 2020 or 2021 regional mortality levels and pre-pandemic mortality. In this group we have [15] for Brazil, [16] and [17] for Italy, [18] for Mexico, [19] for Portugal, [20] for Spain, [21] for Sweden, [22] for Switzerland and [23] for the United States. More thoughtful accounting of the temporal change in mortality via forecasting techniques have been also proposed for estimating excess mortality at regional level. Whereas different countries have been analyzed by [24], country-specific studies have been presented using different methods and aiming to different purposes: [25] for Belgium, [26] for Ecuador, [27] for England and Wales, [28] and [29] for Italy, [30] for Latvia, [31] for Thailand and [32] for United States.

While the value of producing excess mortality measures at a fine geographic scale seems clear and timely, the methodological challenges to estimate them are numerous, and essentially related to the presence of small populations that naturally display high stochastic variation in death counts. On the one hand in this situation possible interpretations of regional differences are necessarily limited. On the other, robust, flexible and fast methods for forecasting mortality levels in the theoretical absence of a pandemic as well as for computing uncertainty around estimates become crucial.

In this study, we propose a novel approach to estimate subnational excess mortality in times of pandemic aiming to cope with all these issues in a unified and clear-cut framework. In details, we use *CP*-splines [33] to project mortality, since this approach exhibits two relevant features when dealing with small area mortality analysis: a large flexibility in modelling diverse demographic scenarios comes along with robustness with respect variation due to small populations at risk. Concerning reliable measures of uncertainty around estimates for the numerous subpopulations at hands, we present a fully analytic procedure with enormous advantages from computational cost and time perspective.

To illustrate this approach, we compute excess mortality in 2020 for the 95 Metropolitan French *départements*, the finest geographical level (NUTS 3) of the classification used by Eurostat. Despite specific outcomes from France, this paper aims to provide a general framework for computing excess mortality and associated uncertainty at subnational level. Given this purpose, R routines [34] are publicly available to replicate our methodology in other countries, and in different historical contexts. See the open source framework repository on this link https://osf.io/zt2c8/?view_only=16ff2a7384c04659bdc39c6a223f2403 [Currently the link is made anonymous for peer review].

## 2 Methods

In this section we give an overview of the methodological aspects behind the computation of excess mortality. Specifically, we measure excess mortality accounting for both historical mortality trends and specific age-pattern in each subnational population. For ease of presentation, we focus on 2020. However, without loss of generality, computation of excess mortality in successive years can be obtained by extending the forecast horizon presented below. Moreover, we compute associated uncertainty in a simple, albeit rigorous, procedure. This allows us to separate main sources of variation before assessing any possible mortality shock in small areas.

For a given subpopulation and sex, we have ***D*** = (*d*_*ij*_) and ***E*** = (*e*_*ij*_), *m* × *n* matrices of deaths and exposures. Number of deaths *d*_*ij*_ at age *i* in year *j* are assumed to be realizations from a Poisson distribution with mean *µ*_*ij*_ *e*_*ij*_ [35], where *µ*_*ij*_ is commonly named force of mortality. In order to compute a theoretical level that would have occurred in the absence of the pandemic, we model observed mortality for *n*_1_ pre-pandemic years and forecast *µ*_*ij*_ for 2020. Among a large number of approaches, we adopt a *CP*-spline model for forecasting mortality in 2020 [33]. This method allows us to simultaneously estimate and forecast mortality within a regression setting with enormous advantages in the computation of uncertainty measures. Let arrange the complete matrices as a column vector, that is, ***d*** = vec(***D***) and ***e*** *=* vec(***E***). Mortality over all ages and years can thus be expressed as the exponential of a linear combination of *B*-splines basis and coefficients:

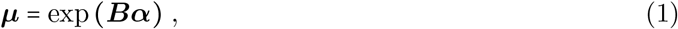

where ***B*** a two-dimensional model matrix that combines *B*-splines over age and years. The associated coefficients ***α*** are penalized to enforce smoothness over both dimensions in order to lie future 2020 mortality within trends estimated from pre-pandemic years. More details can be found in **(author?)** [33].

An important issue when forecasting mortality is the choice of the most appropriate period to apply the forecasting model. Instead of taking all available data or a common starting year for all regions, we optimize the time-window used for each of them. In practice, we apply *CP*-splines with a rolling starting year up to 2010, forecast 2019, and measure the distance between observed and forecast 2019 mortality. Working in a Poisson setting, we opt to measure distance by Deviance [36, p. 34]. The starting year with lowest Deviance value was selected for the final analysis.

A measure of excess mortality for 2020 is defined as the difference between the value of a demographic indicator in a theoretical baseline mortality level and the value for the same indicator obtained from the observed mortality. Whereas the former is obtained by *CP*-splines and it is solely dependent on the estimated coefficients ***α*** in (1), observed death rates ***m***_2020_ **= *d***_2020_/***e***_2020_ are the bases for computing actual level of mortality in the pandemic year.

Besides their estimated values, both observed and forecast values for any mortality measure embody levels of uncertainty that need to be accounted before drawing any conclusions about their change. This consideration is particularly relevant in subnational analyses when relatively small populations at risk are examined. Instead of time-consuming simulation and bootstrap procedures, we develop an analytic construction of the variance associated to both observed and forecast mortality indicators by delta method. With this approach, we identify and disentangle the amount of uncertainty due to either estimated baseline mortality (hereafter “*forecast uncertainty*”) or to observed mortality level in 2020 (hereafter “*Poisson uncertainty*”).

This general framework can be easily adapted to compute excess mortality using a large number of demographic indicators. In the Supplementary Materials A, we provide derivations to obtain these estimates and their associated uncertainty for life expectancy at any given age *x*, age-standardized death rates (SDRs) and plain death toll.

## 3 Data and applications

For illustrative purposes, we present excess mortality measures by losses/gains in life expectancy at age 60 (*e*_60_) in 2020 for 95 *départements* of Metropolitan France, which correspond to the third level of the Nomenclature of Units for Territorial Statistics (NUTS) used by Eurostat. French Human Mortality Database [37] provides annual deaths (***D***) and population at 1st January by single age at death (with an open age interval 95+), sex, and *département* for each year between 1970 and 2020. We define exposures (***E***) as the mean of populations at 1st January for two consecutive years. For comparison, we gather populations and deaths to obtain data for 22 bigger *régions* (NUTS 2 level). Outcomes for this larger geographical level as well as for both sexes are provided in the Supplementary Materials B.

Figure 1 presents losses in male *e*_60_ for a single subpopulation, *Territoire de Belfort*. This *département* was not chosen randomly: strongly affected by the pandemic, it is a relatively small area (about 70,000 men in 2020) and therefore may show more variability in mortality due to smaller sample size.

**Figure 1:**
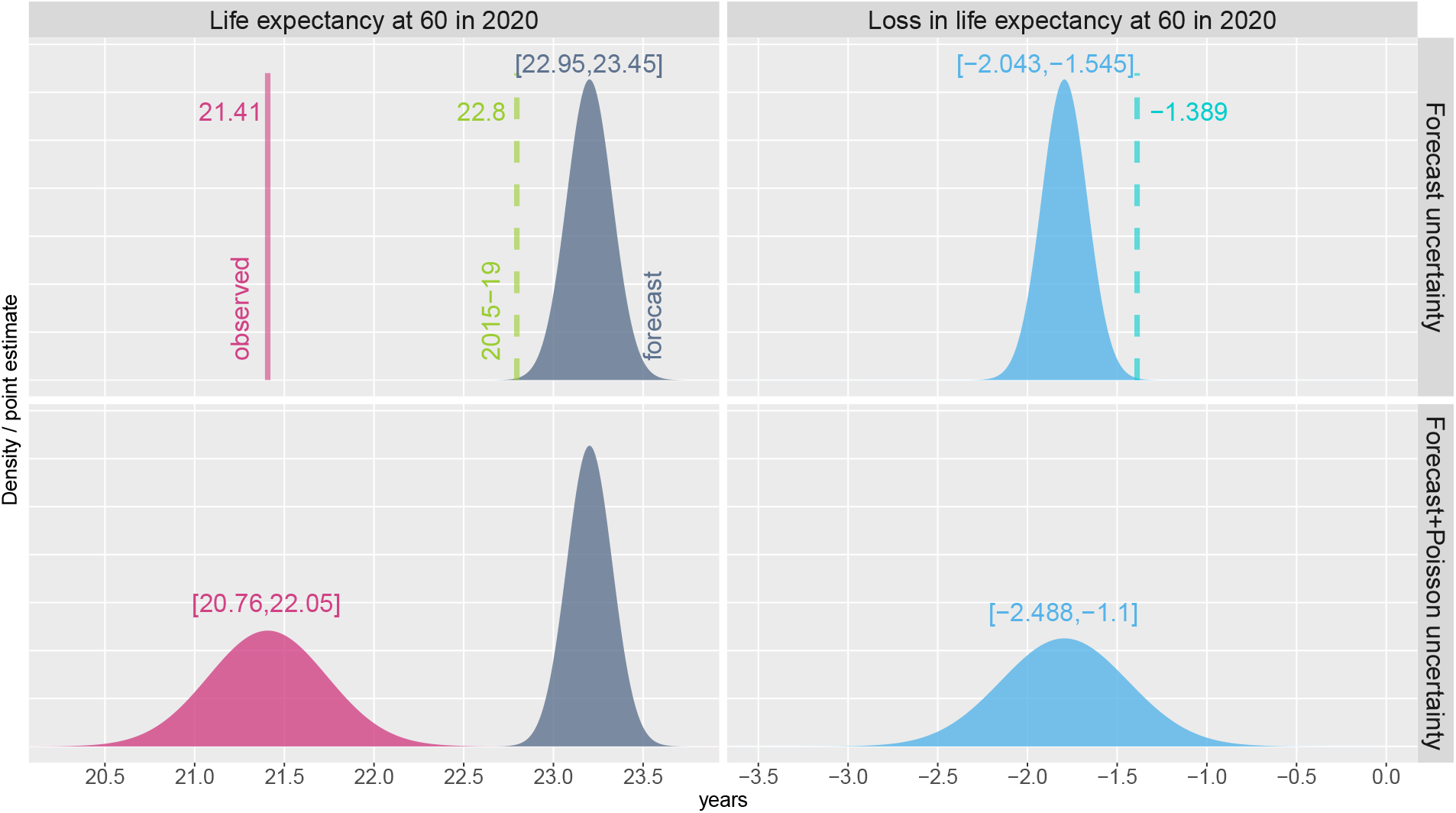
Illustrative figure of sources of uncertainty around excess mortality measure. Life expectancy at 60 (left panels) and associated losses (right panels) for *Territoire de Belfort*, males, 2020. Upper panels: forecast uncertainty is accounted. Lower panel: both forecast and Poisson uncertainty are reported. Texts refer to either point estimates or 95% confidence intervals. Dashed lines depict naive estimation of excess mortality.

The upper panels of Figure 1 reveals how forecast includes uncertainty around estimates. Whereas observed life expectancy at age 60 in 2020 was 21.41 years, our projected value lies between 22.95 and 23.45 years. Consequently, a loss in *e*_60_ is estimated between approximately 1.5 and 2 years. For comparison, Figure 1 presents what we labelled as the naive estimate of excess mortality, commonly used in many recent studies: mortality level from the 5 pre-pandemic years is used as the theoretical baseline level without pandemic. Ignoring decreasing trends in mortality, this approach biases excess mortality estimate downward: loss in *e*_60_ is here only 1.4 years.

The lower part of Figure 1 presents the Poisson uncertainty which is associated to the observed mortality level in 2020. Negligible when the size of the population is large, this source of uncertainty become relevant at regional level. In our example, adding the Poisson uncertainty around our estimates increases the confidence interval by 0.9 year. At last, in 2020 and for *Territoire de Belfort*, we measure a loss in male *e*_60_ between 1.1 and 2.5 years.

In the Supplementary Material B we replicate Figure 1 for *Seine-Saint-Denis*. This *département* was also heavily affected by the pandemic in 2020, but male population is 13 times larger. Total uncertainty is thus much lower (0.8 year) with we estimate a loss in *e*_60_ between 2.4 and 3.2 years.

Figure 2 presents point estimates of losses/gains in *e*_60_ for each *département* along with their 95% confidence interval as well as naive estimates. Both sources of uncertainty, forecast and Poisson, are portrayed. Figure 3 mirrors this information into a French map: point estimates are portrayed and *départements* with non-significant result at the 5% level are highlighted.

**Figure 2:**
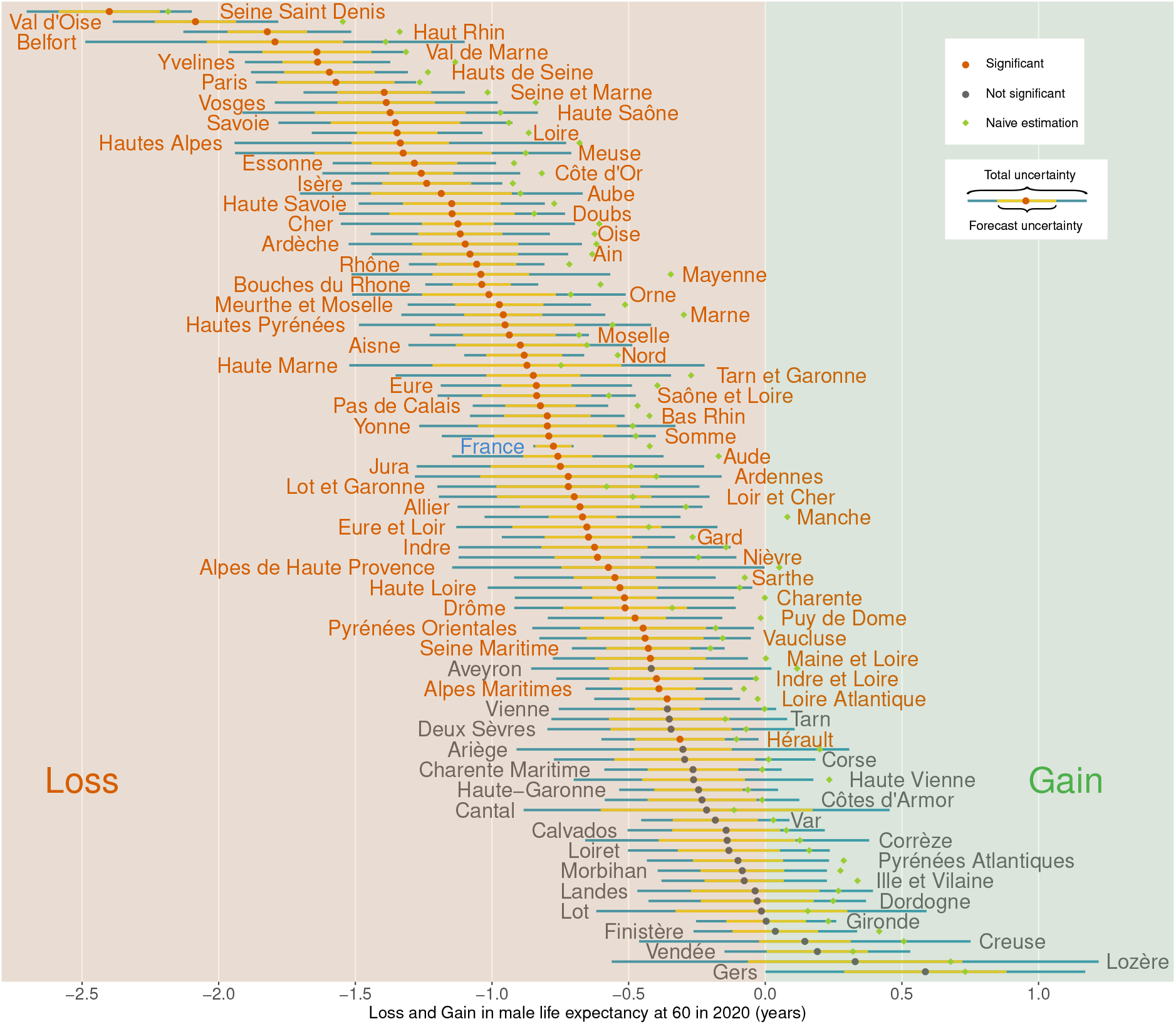
Losses in male life expectancy at age 60 in 2020 for each French *département*. Colors of dots and texts depict express the presence of significant estimates at 95% level, and colors of the horizontal bars represent the two sources of uncertainty. Green dots identify the so-called naive estimation of losses.

**Figure 3:**
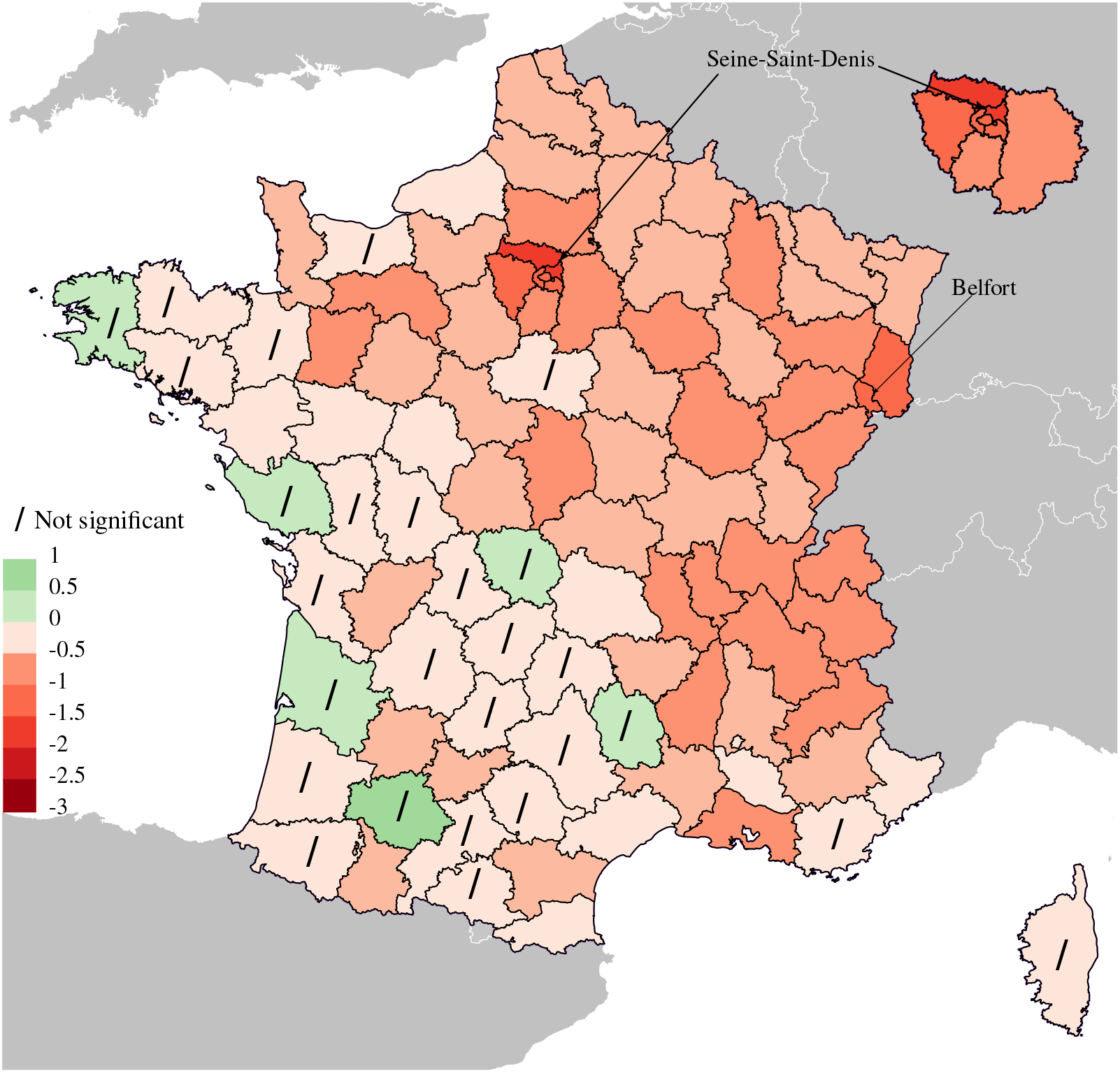
Map of France *département* by losses/gains in male life expectancy at age 60 in 2020. Slash symbol (/) denote areas with loss/gain in *e*_60_ not significant at 5% level. On the upper-right corner zoom of a part of the map referring to Greater Paris (*Ile-de-France*).

In mainland France, we estimate that *e*_60_ has decreased by 0.77 year, whereas the naive estimate is almost two times lower (0.42). The uncertainty around this value is 0.15 year and mostly due to forecast; the Poisson uncertainty almost disappears when we deal with the whole French male population (33 million). Loss in male *e*_60_ remarkably vary across the subpopulations: the maximum loss was in *Seine-Saint-Denis* (2.4 years) whereas the minimum was in *Gers* (gain of 0.6 year). However, 95% confidence intervals around this point estimate is very large (1.2 years) resulting in only a slightly significant gain at the 5% level.

Overall 26 *départements* show not significant estimates at 5% level; only when losses in *e*_60_ rise to about 0.4 year we start detecting significant excess mortality, with the exception of relatively highly populated areas such as *Hérault* and *Loire-Atlantique*. Figure 3 reveals the geography of the pandemic in 2020. Whereas Western France was practically spared by the pandemic, estimates were larger in the East and in the Greater Paris (*Ile-de-France*) region with losses in life expectancy at age 60 about 1 and 1.5 years, respectively.

A central aspect of this paper concerns the importance of measuring uncertainty around excess mortality estimates when dealing with small areas. Figure 4 presents a log-log plot of the amount of uncertainty (measured by the range of the 95% confidence intervals) against population size, i.e. we portray the proportional change in uncertainty in response to a proportional change in population size. In order to broaden the view, we portray both *départements* (NUTS 3) and *régions* (NUTS 2) and, along with the total uncertainty (in green), we differentiate uncertainty coming from the forecast procedure (in purple) and from Poisson on observed data (in orange).

**Figure 4:**
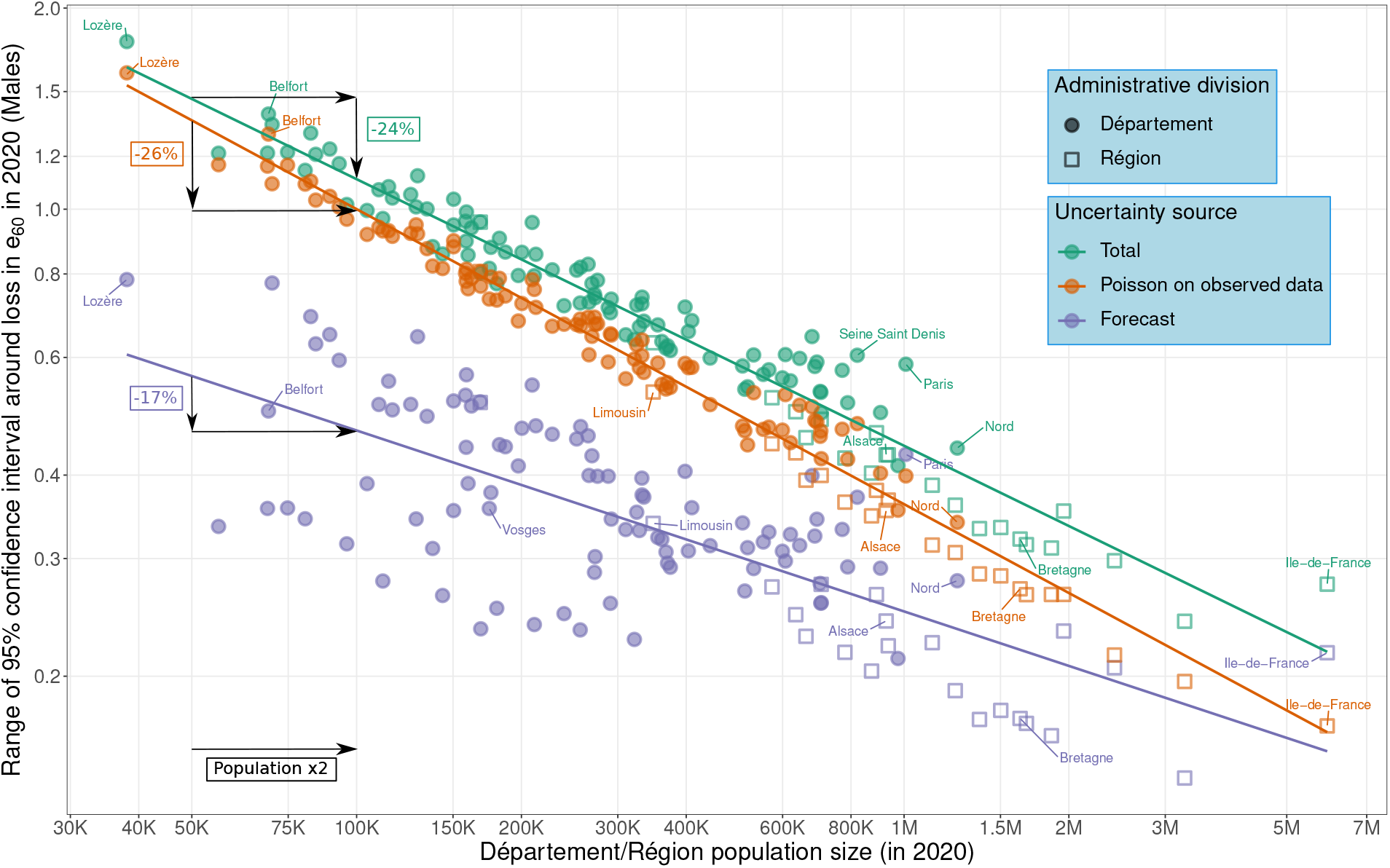
Log-log plot of the amount of uncertainty against population size by source of uncertainty: Total, associated to the Forecast process and due to Poisson randomness in observed data. Uncertainty is measured by the width of the 95% confidence intervals around estimated loss in male *e*_60_. Values for both *départements* (NUTS 3) and *régions* (NUTS 2) are depicted. Linear fits are provided for illustrative purposes and for obtaining an approximated value of the elasticity associated to each source of uncertainty.

With a linear fit to both departmental and regional values, we are able to estimate elasticity associated to source-specific uncertainty and gauge the percent change in uncertainty for a doubling in population size. In general, we estimate that uncertainty decreases by 24% when the population doubles. This value combines two sources of uncertainty that decrease at an unequal pace when population grows. Whereas Poisson uncertainty is higher than the forecast uncertainty for almost all subnational populations, this source of uncertainty decreases at faster pace (26%) than the forecast uncertainty (17%) when the population doubles.

We can also read Figure 4 from an alternative perspective. When excess mortality is measured by *e*_60_, a loss larger than 0.75 years would be necessary to have a significant estimate at 95% level if the population size is 50,000. This value decrease with larger populations: in a region with 200,000 (one million) men, a loss in *e*_60_ equal to 0.4 (0.2) would be required to obtain a significant excess mortality.

## 4 Discussion

Assessing excess mortality during a pandemic such as the recent Covid-19 is as crucial from an health policy perspective as challenge from a methodological view. Moreover these challenges increase when we deal with excess mortality estimation at subnational level. Specifically we face two main issues.

First, computation of mortality levels that would have been observed without pandemic need to be estimated. In other words, forecasting methods are necessary to extrapolate temporal mortality variations. In this paper, we use *CP*-splines [33] and illustrate our approach with a reproducible example on French *départements*. If data on deaths and exposure population are available by age and year, the proposed approach is flexible to adapt to a large variety of current, and historical, scenarios and robust for dealing with very small populations.

Second, uncertainty become very high when one deals with low populated areas. In this paper, we compute uncertainty around point estimates, and disentangle between uncertainty due to the forecasting process and inherent uncertainty due to Poisson random nature of observed mortality data. We show that overall uncertainty in excess mortality decreases by 24% when population doubles, though Poisson uncertainty tends to decrease more rapidly when population grows. Consequently, whereas for large populations we can safely disregard uncertainty in the observed data and account only for the forecasting errors, when dealing with excess mortality in small areas, the Poisson component in the uncertainty must be take into account before drawing any conclusions.

A possible way for reducing uncertainty, namely the Poisson component, is to either gather populations spatially by aggregating smaller administrative divisions into larger ones, or estimating excess mortality for both sexes. Still, these choices must be made with caution since associated outcomes might hide strong heterogeneity.

To illustrate these concepts Figure 5 shows densities of losses in male *e*_60_ for two specific French *régions* (lower panel) and associated lower level administrative divisions, *départements* (upper panel). In this example, *Ile-de-France* or Grater Paris is the *région* that suffered from the highest loss in life expectancy at age 60 with a 95% confidence interval loss in *e*_60_ of [1.57 − 1.85] . Still, this result hides large heterogeneity between the least and the harshest hit *département* in this *région*: *Esonne* with [0.98 − 1.68] and *Seine-Saint-Denis* with [2.10 − 2.70]. It is noteworthy that mortality level in *Seine-Saint-Denis* was already among the highest in France before 2020. Concealing its further deterioration caused by the pandemic for the sake of reducing uncertainty by a spatial aggregation will thus be an inappropriate procedure from an health policy perspective. On the contrary, estimates for *Bretagne* do not mask large spatial heterogeneity and aggregation within this region considerably decreases size of the confidence interval around the excess mortality estimates without much loss of information.

**Figure 5:**
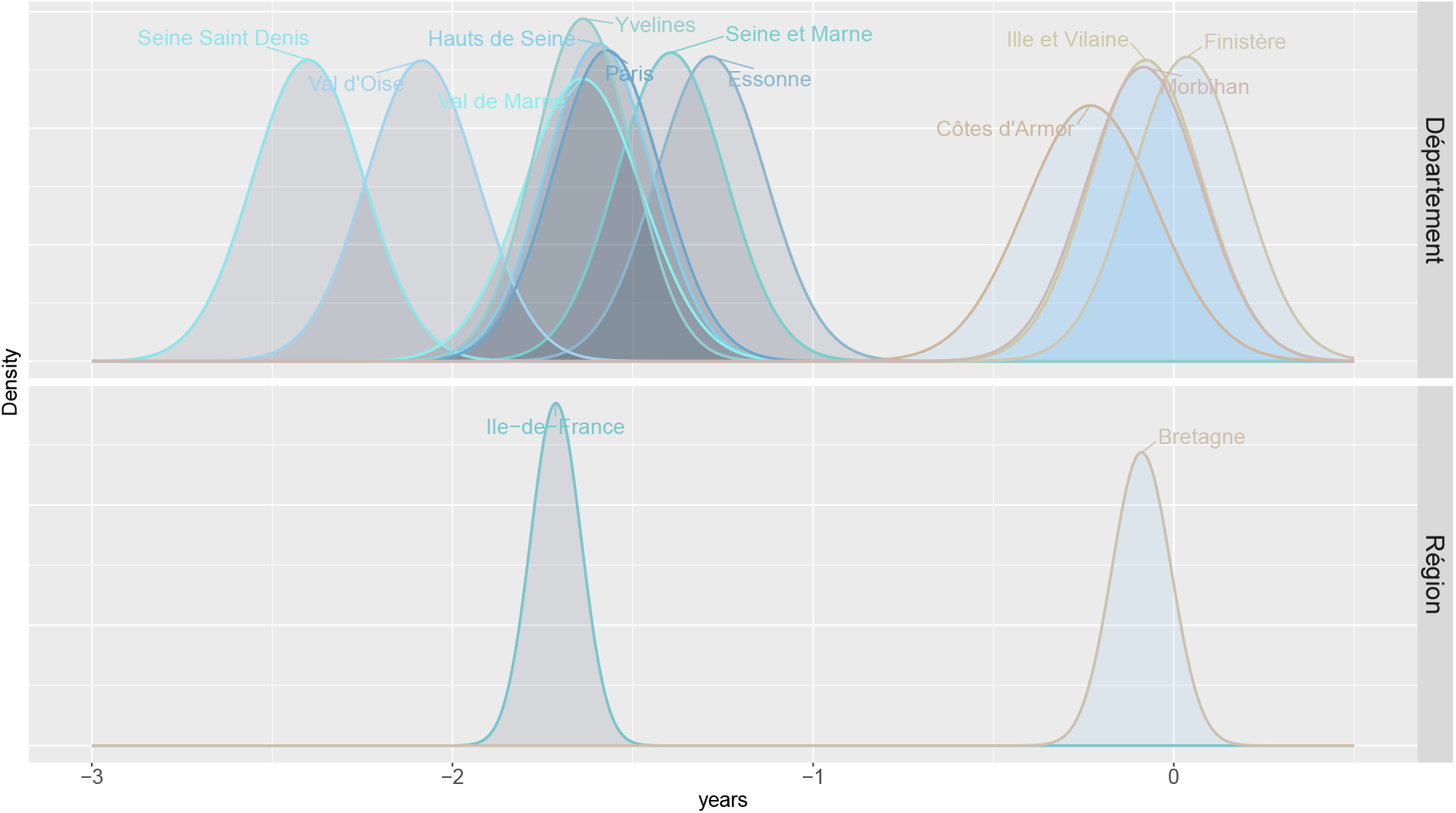
Illustrative figure on the effects of spatial aggregation in excess mortality estimation. Densities of losses/gains in life expectancy at 60 for two NUTS 2 populations (*Ile-de-France* and *Bretagne*, lower panel) and their associated NUTS 3 populations (upper panel).

An alternative way for reducing uncertainty keeping the same administrative division would be to combine men and women. On the one hand, practically doubling population sizes, this strategy would reduce the width of associated 95% confidence intervals by 24%. On the other estimates for both sexes will surely hide heterogeneity between men and women in a context in which literature has revealed large sex differences in Covid-19 morbidity and mortality. See, among others, [38; 39; 40; 41; 42]. For completeness, Supplementary Materials B present excess mortality estimates measured by *e*_60_ for both sexes combined as well as for NUTS 2 regions.

In summary, unlike most of the previous methods proposed to estimate excess mortality, our approach allows to cope with all issues akin with small populations. Thanks to its robustness, flexibility and low computational cost, we envisage its wider use for mapping the impact of Covid-19 at the international level. We also encourage national statistical offices to continue and expedite publication of mortality data at regional level that, coupled with our available routines, would facilitate a more accurate and timely assessment of the burden of any ongoing pandemic.

## Data Availability

All data produced are available online at :
https://osf.io/zt2c8/?view_only=16ff2a7384c04659bdc39c6a223f2403

