## Supplementary Material for "Estimating Subnational Excess Mortality in Times of Pandemic. An application to French *départements* in 2020"

### Estimating Subnational Excess Mortality in Times of Pandemic.
### Supplementary material

November 29, 2022

#### Introduction

In this supplementary material, Section A presents derivations for computing, from $CP$-spline estimated coefficients and observed mortality rates, excess mortality measured by different demographic indicators. Additional figures for French mortality data are given in Section B.

#### A Methods to compute excess mortality

When measuring excess mortality in our setting, there are concepts which are common to all demographic indicator employed. On the one hand, baseline mortality that would have been observed in the absence of a pandemic is fully represented by the coefficients vector $\boldsymbol{\alpha}$ estimated via $CP$-splines (see equation (1) in the manuscript). On the other, level of mortality observed in 2020 is described by the series of coefficients $\boldsymbol{m}_{2020}$.

We can generally measure excess mortality by the difference between a demographic indicator estimated by $\boldsymbol{\alpha}$ and $\boldsymbol{m}_{2020}$:

$$\delta_f = f^F(\boldsymbol{\alpha}) - f^O(\boldsymbol{m}_{2020}) \tag{A.1}$$

where $f^F(\cdot)$ and $f^O(\cdot)$ denote a generic function for forecast and observed mortality level, respectively.

Variances associated to both forecast and observed mortality level are given by for a generic function:

$$\begin{aligned} \mathbb{V}(f^F(\boldsymbol{\alpha})) &= \nabla f^F(\boldsymbol{\alpha}) \operatorname{cov}(\boldsymbol{\alpha}) \nabla f^F(\boldsymbol{\alpha})' \\ \mathbb{V}(f^O(\boldsymbol{m}_{2020})) &= \nabla f^O(\boldsymbol{m}_{2020}) \operatorname{cov}(\boldsymbol{m}_{2020}) \nabla f^O(\boldsymbol{m}_{2020})', \end{aligned} \tag{A.2}$$

where symbol $\nabla$ denotes the vector differential operator, i.e. the partial derivatives of life expectancy with respect to either $\boldsymbol{\alpha}$ or $\boldsymbol{m}_{2020}$.

Since observed 2020 mortality is described by the associated death rates, the covariance matrix is simply a diagonal matrix of the inverse of the observed deaths: $\operatorname{cov}(\boldsymbol{m}_{2020}) = \texttt{diag}(1/\boldsymbol{d}_{2020})$.

Regarding the forecast mortality, we need to account for the full covariance structure in the $CP$-spline model. Unlike in the original paper, we borrowed its structure from regression theory and $\text{cov}(\boldsymbol{\alpha})$ will be computed from estimated values. See Eilers and Marx (2021, p. 32-33). It is of note here a seemingly paradoxical behaviour, often overlooked in the literature and independent of the employed forecasting approach: uncertainty associated to observed mortality is expected to be larger than variability around forecast mortality in 2020. Whereas the former depends on observed mortality for a specific independent year, forecast mortality in 2020 is the product of a model which incorporates information over both age and time and hence it reduces variability around the fitted values, in spite of dealing with a future, albeit single, year.

Assuming $f^F(\boldsymbol{\alpha})$ and $f^O(\boldsymbol{m}_{2020})$ as independent and asymptotically normal distributed, we can easily compute the variance for the excess mortality measure in (A.1) as the sum of the variances presented in (A.2). Moreover, valuing more clarity than conciseness, in the following we will present equations without proceeding with utmost simplifications.

#### A.1 Life expectancy

Life expectancy is defined as the average length of life of a newborn if current age specific mortality rates in that population applied in the future. Widely used to assess health status of a population, it can be computed at any given age.

For illustrative purposes, we start with the most common life expectancy at birth $e_0$ and, in matrix notation, generic (A.1) becomes:

$$\delta_{e_0} = e_0^F(\boldsymbol{\alpha}) - e_0^O(\boldsymbol{m}_{2020}) = \mathbf{1}'_m \left[\exp(\boldsymbol{C}\,\boldsymbol{L}\,\boldsymbol{\mu}) - \exp(\boldsymbol{C}\,\boldsymbol{m}_{2020})\right] , \qquad \text{(A.3)}$$

where $\boldsymbol{C}$ is a $m \times m$ lower-triangular matrix with -1 entries for computing the cumulative summation of mortality, e.g. $\boldsymbol{C}\boldsymbol{m}_{2020}$ corresponds to the discrete counterpart of minus the cumulative observed hazard function in 2020. The matrix $\boldsymbol{L}$ is constructed in order to select 2020 forecast mortality from equation (1) in the paper. The $m \times 1$ matrix of 1s, $\mathbf{1}_m$, serves to sum up the exponential of minus the cumulative hazards over all ages.

Partial derivatives of life expectancy necessary for computing variances of excess mortality as in (A.2) are given by

$$\begin{aligned} \nabla e_0^F(\boldsymbol{\alpha}) &= \mathbf{1}'_m\, \texttt{diag}(\exp(\boldsymbol{C}\,\boldsymbol{L}\,\boldsymbol{\mu}))\,\boldsymbol{C}\,\boldsymbol{L}\,\texttt{diag}(\boldsymbol{\mu})\,\boldsymbol{B} \\ \nabla e_0^O(\boldsymbol{m}_{2020}) &= \mathbf{1}'_m\, \texttt{diag}(\exp(\boldsymbol{C}\,\boldsymbol{m}_{2020}))\,\boldsymbol{C}\,\texttt{diag}(\boldsymbol{m}_{2020})\,. \end{aligned} \qquad \text{(A.4)}$$

Life expectancy at any age $x$ can be obtained by modifying vector $1_m$ and matrix $\boldsymbol{C}$, i.e. replace with zeros rows/columns corresponding to ages smaller than $x$. Similar arguments can be applied to age-standardized mortality rates and plain death toll presented below.

#### A.2 Age-standardized death rate

An age-standardized mortality rate (SDR) is the crude death rates of a population of size $\kappa$ adjusted to a standard age distribution, $\boldsymbol{p}$, and it is calculated as a weighted average of the age-specific mortality rates. In matrix notation excess mortality measured by SDR ia given by

$$\delta_{SDR} = SDR^F(\boldsymbol{\alpha}) - SDR^O(\boldsymbol{m}_{2020}) = \kappa * (\boldsymbol{p} * \mathbf{1}_m)'(\boldsymbol{L}\boldsymbol{\mu} - \boldsymbol{m}_{2020})\,, \qquad \text{(A.5)}$$

where  denote element-wise product. In the available routines, European Population Standard of 2013 (European Commission, 2013) is provided for $\boldsymbol{p}$.

Partial derivatives of SDR for computing variances of excess mortality are given by

$$\begin{aligned} \nabla SDR^{F}(\boldsymbol{\alpha}) &= \kappa * (\boldsymbol{B}'\,(\boldsymbol{L}'\,(\boldsymbol{p} * \mathbf{1}_m) * \boldsymbol{\mu}))' \\ \nabla SDR^{O}(\boldsymbol{m}_{2020}) &= \kappa * (\boldsymbol{p} * \mathbf{1}_m * \boldsymbol{m}_{2020})' \,. \end{aligned} \tag{A.6}$$

#### A.3 Death toll

Excess mortality can be simply computed as the difference between the total death toll that would have been observed in the absence of a pandemic and the actual death toll reported in the year of pandemic (here 2020). Whereas life expectancy and SDR are age-standardized measures, total number of deaths during the pandemic will depend upon structure and size of the population. However, this measure is very easy even for layman to understand and use and it can provide a direct assessment of the actual burden of the pandemic in a given region.

Computed as follows:

$$\delta_{DT} = DT^{F}(\boldsymbol{\alpha}) - DT^{O}(\boldsymbol{m}_{2020}) = \mathbf{1}_m\,[\boldsymbol{L}(\boldsymbol{e} * \boldsymbol{\mu}) - \boldsymbol{d}_{2020}] \,, \tag{A.7}$$

the partial derivatives of this measure for both baseline and observed mortality are given by

$$\begin{aligned} \nabla DT^{F}(\boldsymbol{\alpha}) &= (\boldsymbol{B}'\,(\boldsymbol{L}'\,\mathbf{1}_m * \boldsymbol{e} * \boldsymbol{\mu}))' \\ \nabla DT^{O}(\boldsymbol{m}_{2020}) &= (\mathbf{1}_m * \boldsymbol{d}_{2020})' \,. \end{aligned} \tag{A.8}$$

### B Additional figures

In this section we present few additional figures on our application to French mortality data.

Figure B.1 present the same outcomes than Figure 1 in the paper. The difference concerns the size of the portrayed *département*: *Seine-Saint-Denis* is a much larger population than *Territoire de Belfort*. However, inclusion of both sources of uncertainty in estimating excess mortality is also evident here.

Figure B.2 shows excess mortality measured by life expectancy at age 60 for all French *départements* (NUTS 3) both both sexes combined. As in the related figure in the manuscript for males only (Fig. 2) we plot 95% confidence intervals obtained by including both forecast and Poisson sources of uncertainty. An clear reduction in the uncertainty around the estimated excess mortality is clear when both sexes are combined. However, loss in terms of precision needs to be accounted, loss which is relevant in case of pandemics with large sex differences in morbidity and mortality.

Figure B.3 presents losses in life expectancy at 60 in 2020 for males for each of the 22 French *régions* (NUTS 2). Likewise the previous figures 95% confidence intervals as sources of uncertainty are plotted.

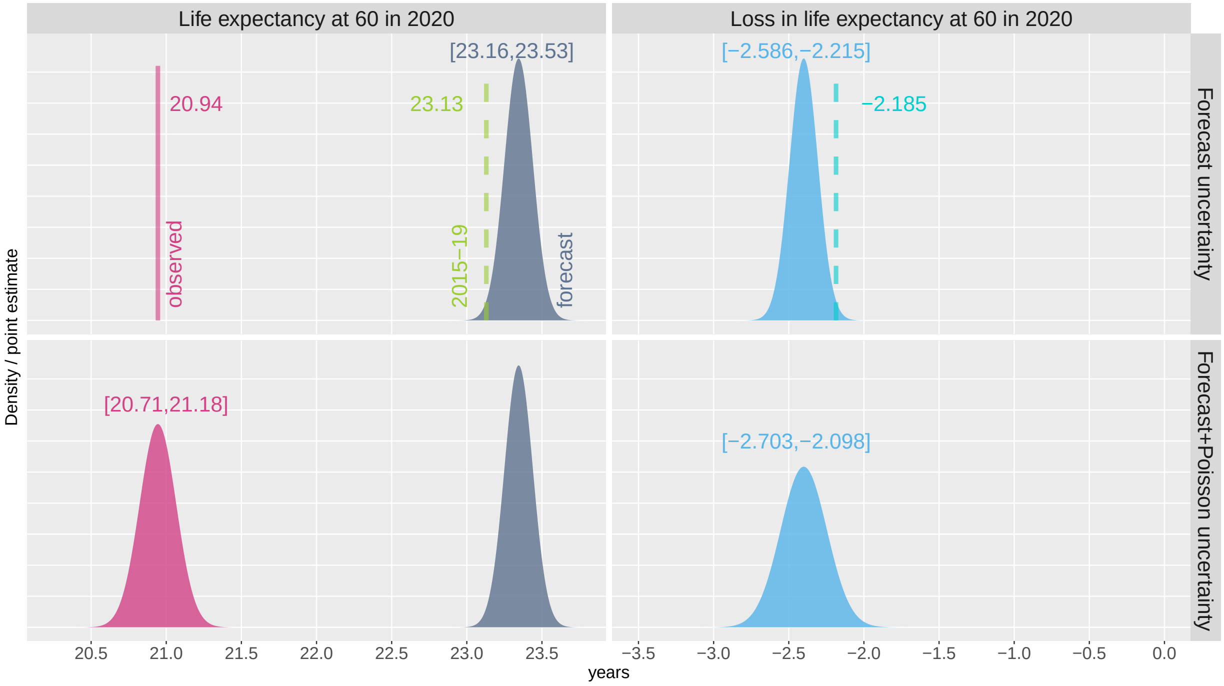


Figure B.1: Illustrative figure of sources of uncertainty around excess mortality measure. Life expectancy at 60 (left panels) and associated losses (right panels) for *Seine-Saint-Denis*, males, 2020. Upper panels: forecast uncertainty is accounted. Lower panel: both forecast and Poisson uncertainty are reported. Texts refer to either point estimates or 95% confidence intervals. Dashed lines depict naive estimation of excess mortality.

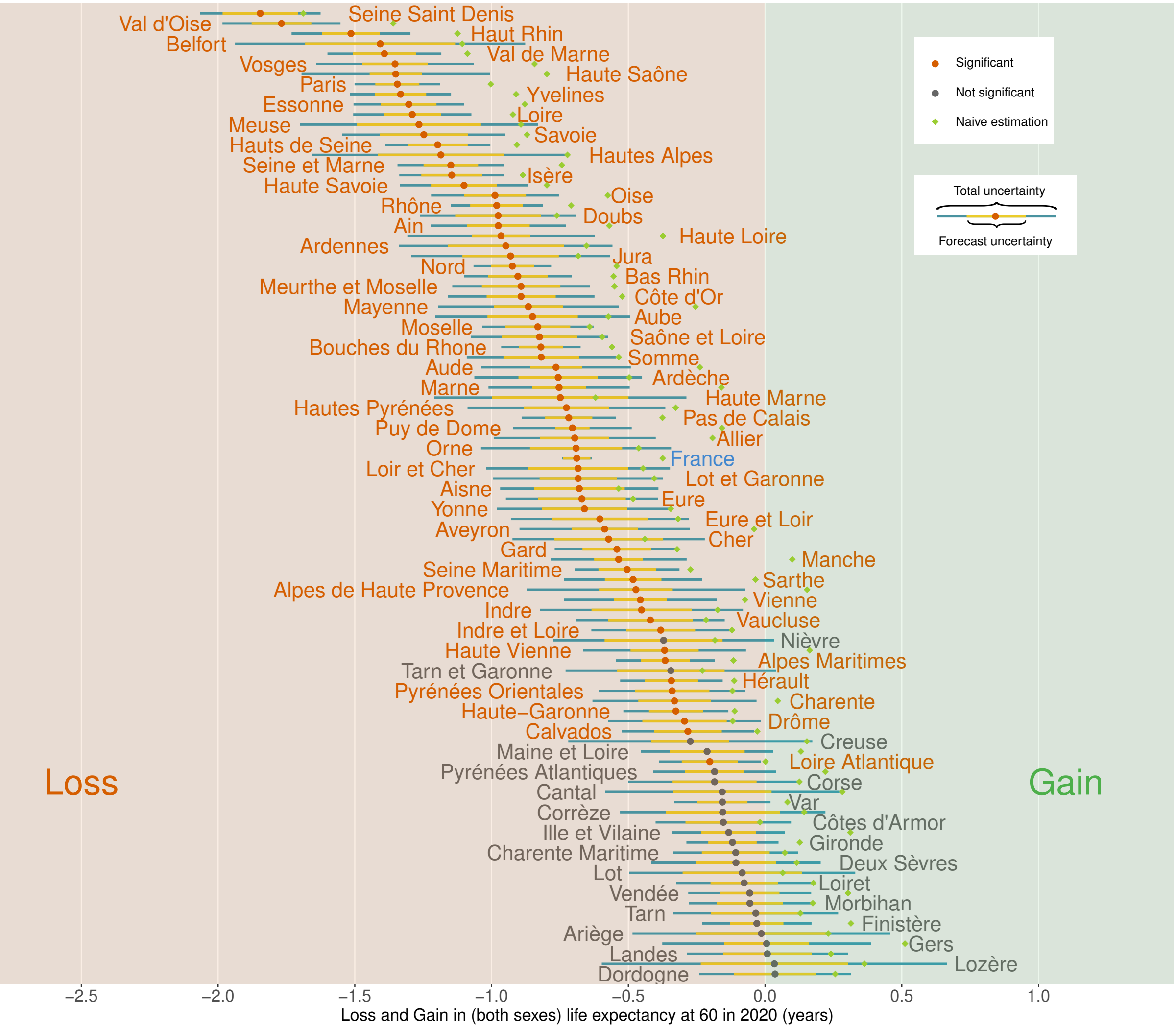


Figure B.2: Losses in life expectancy at age 60 in 2020 for each French *département*, both sexes combined. Colors of dots and texts express the presence of significant estimates at 95% level, and colors of the horizontal bars represent the two sources of uncertainty. Green dots identify the so-called naive estimation of losses.

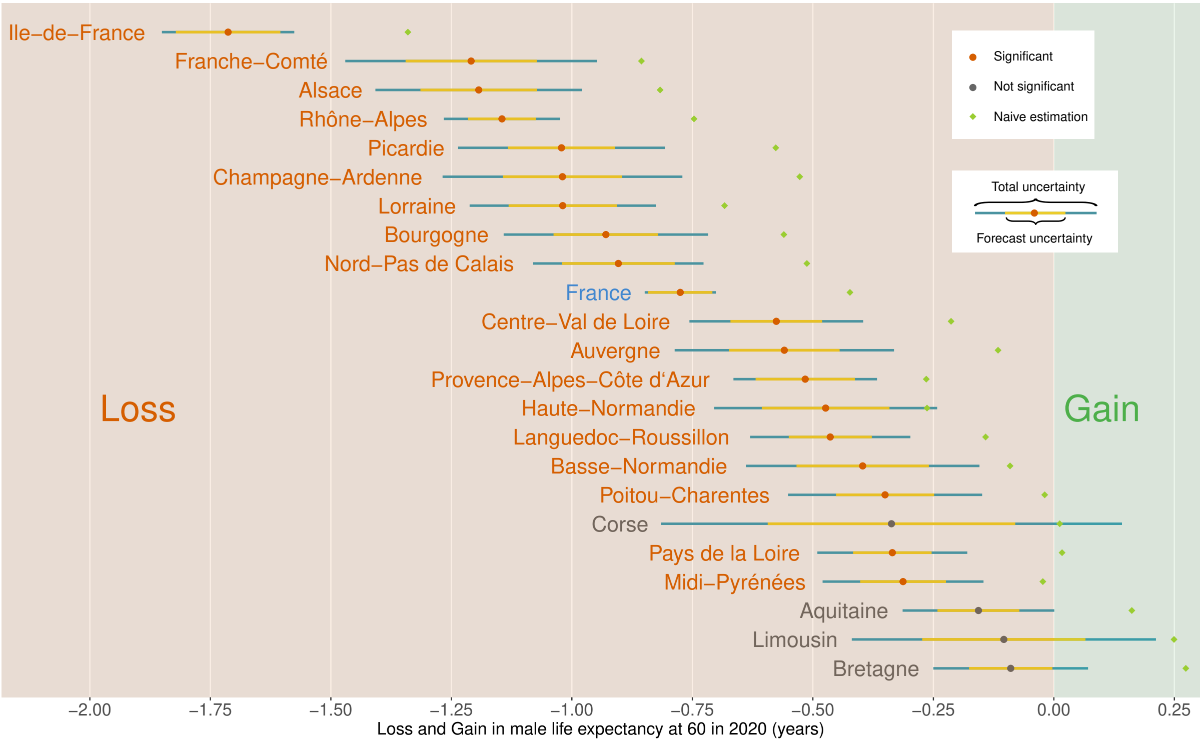


Figure B.3: Losses in male life expectancy at age 60 in 2020 for each French *régions*. Colors of dots and texts express the presence of significant estimates at 95% level, and colors of the horizontal bars represent the two sources of uncertainty.
